# Population-Based Model of the Fraction of Incidental COVID-19 Hospitalizations During the Omicron BA.1 Wave in the United States

**DOI:** 10.1101/2022.01.22.22269700

**Authors:** Jeffrey E. Harris

## Abstract

**Background:** Some reports have suggested that as many as one-half of all hospital inpatients identified as COVID-19-positive during the Omicron BA.1 variant-driven wave were incidental cases admitted primarily for reasons other than their viral infections. To date, however, there are no prospective longitudinal studies of a representative panel of hospitals based on pre-established criteria for determining whether a patient was in fact admitted as a result of the disease.

**Materials and Methods:** To fill this gap, we developed a formula to estimate the fraction of incidental COVID-19 hospitalizations that relies upon measurable, population-based parameters. We applied our approach to a longitudinal panel of 164 counties throughout the United States, covering a 4-week interval ending in the first week of January 2022.

**Results:** Within this panel, we estimated that COVID-19 incidence was rising exponentially at a rate of 9.34% per day (95% CI, 8.93-9.87). Assuming that only one-quarter of all Omicron BA.1 infections had been reported by public authorities, we further estimated the aggregate prevalence of active SARS-CoV-2 infection during the first week of January to be 3.45%. During the same week, among 250 high-COVID-volume hospitals within our 164-county panel, an estimated 1 in 4 inpatients was COVID-positive. Based upon these estimates, we computed that 10.6% of such COVID-19-positive hospitalized patients were incidental infections. Across individual counties, the median fraction of incidental COVID-19 hospitalizations was 9.5%, with an interquartile range of 6.7 to 12.7%.

**Conclusion:** Incidental COVID-19 infections appear to have been a nontrivial fraction of all COVID-19-positive hospitalized patients during the Omicron BA.1 wave. In the aggregate, however, the burden of patients admitted for complications of their viral infections was far greater.

## Introduction

During the massive wave of Omicron variant-driven SARS-CoV-2 infections in the United States during the winter of 2021–2022, total hospitalizations among infected persons reached 161,876 on January 19, 2022, a nationwide census substantially exceeding the peaks recorded during the previous winter of 2020–2021 and the Delta variant-driven wave of July– October 2021 [1]. Yet some observers cautioned that more than half of all hospital inpatients identified as COVID-19-positive during the Omicron BA.1 surge were so-called incidental cases admitted primarily for reasons other than their viral infections [2–4]. Such estimates, if accurate, would imply that raw counts of COVID-19-associated hospitalizations substantially overstated not only the typical severity of Omicron BA.1 infection but also the overall impact of the surge on healthcare resource utilization.

The notion that some patients hospitalized for entirely unrelated reasons happen to be SARS-CoV-2-positive appears entirely reasonable, especially in view of the substantial fraction of asymptomatic cases among Omicron BA.1 infections [5]. The real problem is putting an accurate bound on the proportion of such incidental hospitalizations. To that end, the ideal study design would be to prospectively follow a representative, longitudinal cohort of individuals, evaluating each hospitalization according to pre-established, objective criteria for determining whether each SARS-CoV-2-positive patient was in fact admitted as a result of his infection.

At issue here are more than technical matters of disease classification, but also fundamental questions of ultimate causation [6]. Consider a patient with hypertensive heart disease who is hospitalized for a tachyarrhythmia due to the mild hypoxemia he suffered from COVID-19. Or consider a young adult recently infected with SARS-CoV-2 who continues to drive while suffering from viremia-induced headache, fever, body aches, dizziness and fatigue and, as a result, gets into a major car crash. The former patient’s admission diagnoses might include such proximate causes as cardiomegaly and atrial fibrillation, while the latter patient’s admission diagnoses might list acute brain injury, spleen laceration and a pelvic fracture. But the ultimate cause in both cases was COVID-19.

Unfortunately, the few documented research studies to date have not achieved this methodological ideal. A tabulation of 2,688 COVID-19 positive hospitalized patients posted by the Massachusetts Department of Public Health during the Omicron BA.1 wave [7] designated only 1,337 (48.8%) as hospitalized due to their SARS-CoV-2 infections. However, these non-incidental admissions were restricted to those patients who had been administered dexamethasone, a treatment principally indicated for severe cases requiring high-flow oxygen but contraindicated in some hospitalized COVID-19 patients [8,9].

A retrospective review of electronic health records of 1,123 patients found to be polymerase chain reaction (PCR) positive from 7 days before to 14 days after admission to one of four U.S. hospital systems [10] did not cover the period of the Omicron BA.1 surge. Among the 292 cases (or 26%) retrospectively classified as incidental, about two-thirds involved such diagnoses as acute kidney injury, diabetes, anemia, thrombocytopenia and acute respiratory failure, which could well have been precipitated or exacerbated by SARS-CoV-2 infection [11–13].

A smaller study of PCR-positive admissions to a tertiary-care hospital in Rotterdam, The Netherlands, during the Omicron BA.1 wave [14] reported that 46 (30.5%) of 151 adults and 8 (38.1%) of 21 pediatric patients had no COVID-19 symptoms or only mild symptoms that did not require any treatment. Once again, there is a genuine question whether treatment of self-reported symptoms is the appropriate criterion for determining whether SARS-CoV-2 causally contributed to a patient’s hospital admission.

In another retrospective review of a large database of electronic health records from 960 U.S. hospitals, the estimated proportions of incidental COVID-19 admissions during the Omicron BA.1 surge in January 2022 were: 25% among elderly patients aged 65 years or more, 45% among adults aged 19-64 years, and 65% among pediatric patients aged 18 years or younger [15]. Patients were classified as incidental if they tested positive on admission but had no record of treatment for COVID-19 or a related respiratory condition. The search criteria did not include nirmatrelvir-ritonavir (Paxlovid™), an oral antiviral combination that entered into widespread use upon its emergency approval by the U.S. Food and Drug Administration in December 2021 [16]. As with other retrospective searches of electronic health records, this study did squarely confront the fundamental issue of causation.

The aim of the present study is to inquire whether the gap in our understanding of the extent of incidental hospitalizations during the Omicron BA.1 wave in the United States can be filled with the tools of integrative epidemiology, an emerging discipline that combines diverse, often unconventional data sources to attack knotty problems of disease causation [17,18].

To that end, we developed a formula to estimate the fraction of incidental COVID-19 hospitalizations that relies upon objectively measurable, population-based parameters. We applied this approach to a longitudinal panel of 164 counties throughout the United States, which contained 250 high-COVID-volume hospitals. Our analysis covered the 4-week interval ending in the first week of January 2022, during which time the Omicron BA.1 variant of SARS-CoV-2 was far and away the dominant strain [19].

## Materials and Methods

### Defining the Fraction of Incidental COVID Hospitalizations, π

We developed a model to formally define the fraction of incidental COVID-19 hospitalizations and to express this fraction as a function of observable quantities. To that end, consider a closed population at a particular point in time. We refer to any individual within this population who has a detectable SARS-CoV-2 infection at that time as a *COVID-19 case*. This definition does not require that the individual’s infection has in fact been detected, either via a positive test or any other means. Nor does it require that the individual has any symptoms of COVID-19.

Let *q* (where 1 > *q* > 0) denote the proportion of all COVID-19 cases that are hospitalized *because* of their illness. For shorthand, we refer to these as the *severe* cases. We refer to all other COVID-19 cases, whether they are hospitalized or not, as *non-severe*. While the severe cases are, by definition, hospitalized because of their COVID-19 illness, some of the remaining non-severe cases will also be hospitalized for non-COVID reasons. We refer to the latter group as *incidental COVID-19 hospitalizations*. Let *g* denote the proportion of non-severe cases that is hospitalized, where 1 > *g* > 0. Then, by Bayes’ rule, the proportion of all COVID-19 hospitalizations that are incidental is:

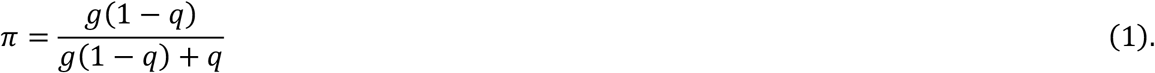

We refer to *π* as the *fraction of incidental COVID-19 hospitalizations*. We see that *π* is increasing in *g* and decreasing in *q*.

### Rendering π, the Fraction of Incidental Hospitalizations, as a Function of Observables

Ideally, one would estimate the quantity *q* in equation (1) by longitudinally following a closed cohort of individuals as they contracted COVID-19 and then observing who was hospitalized because of their illness. Within this longitudinal cohort, we could also estimate *g* by tracking who was hospitalized for other reasons. In the absence of such a longitudinal study, we can still proceed provided we make a key simplifying assumption, namely, that those individuals without COVID-19 have the same probability of hospitalization as those with non-severe COVID-19. We examine the validity of this key assumption later in the Discussion.

Let *p* (where 1 > *p* > 0) denote the proportion of individuals in the entire population who are COVID-19 cases. We refer to *p* as the *prevalence of COVID-19*. Now focus more sharply on just those individuals who are hospitalized. We test every hospitalized patient to determine who is COVID-19-positive and who is COVID-19-negative. Let *c* (where 1 > *c* > 0) denote the fraction of all hospitalized individuals who are COVID-19-positive. In the Supplement, we derive the following expression for *π*:

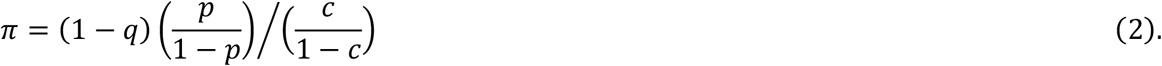

In equation (2), *π* is now a function of the prevalence *p* of COVID-19 in the population, the fraction *c* of all hospitalized patients who are COVID-19-positive, and the proportion *q* of COVID-19-positive individuals requiring hospitalization because of their disease. Table 1 below summarizes these points.

**Table 1.** Component Parameters in the Derivation of the Fraction of Incidental Hospitalizations, *π*.

| **Table 1. Component Parameters in the Derivation of the Fraction of Incidental Hospitalizations, $\pi$** |  |
| --- | --- |
| *Symbol* | *Definition* |
| $p$ | Prevalence of COVID-19 |
| $c$ | Fraction of all hospitalized patients who are COVID-19-positive |
| $q$ | Proportion of all infected individuals who are hospitalized because of their COVID-19 illness |
| $\pi$ | Fraction of incidental hospitalizations, $\pi = (1 - q) \left( \frac{p}{1 - p} \right) / \left( \frac{c}{1 - c} \right)$ |

### Rendering p, the Prevalence of COVID-19, as a Function of Observables

To estimate the prevalence *p*, we would ordinarily rely upon the classic formula in epidemiology, that is, prevalence equals the incidence of SARS-CoV-2 infection per unit time multiplied by the average duration of infection. The difficulty with this formula is that it applies only to a population with a stable incidence rate, and that is certainly not the case here [20].

Let *h*(*t*) denote the incidence rate of SARS-CoV-2 infection at time *t* ≥ 0. We assume that incidence is growing exponentially, that is, 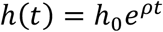, where *h*_0_, *ρ* > 0. Let the duration *s* > 0 of infection have an exponential distribution with mean 1/*θ*, where *θ* > 0. In the Supplement, we show that the prevalence of infection can be approximated as:

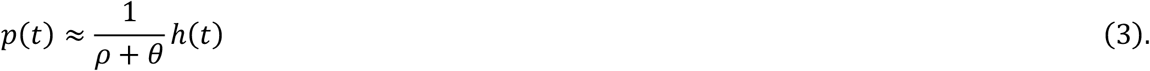

When the incidence *h* is stable (that is, *ρ* = 0), this formula collapses to the classic result *p*(*t*) = *h*(*t*)/*θ*. To estimate the prevalence *p* during an exponentially growing epidemic, we therefore need data on the incidence *h*, the growth rate *ρ* of infection and the mean duration 1/*θ* of infection.

It is widely acknowledged that COVID-19 cases have been and continue to be significantly underreported [21–24]. Accordingly, to estimate the incidence *h* from data on reported cases, we follow the usual approach of incorporating an under-ascertainment factor into our analysis [25]. Let *r*(*t*) denote the *reported* incidence of COVID-19 and let *f*(*t*) > 0 denote the fraction of COVID cases that are reported at time *t*. Then we have:

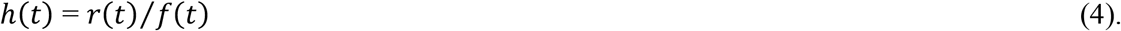

Accordingly, to estimate *p*, we need data on the parameters *r, ρ, f, h*, and *θ*, as indicated in Table 2. For clarity, we have dropped the time argument *t* from the functions *h, r*, and *f*.

**Table 2.**
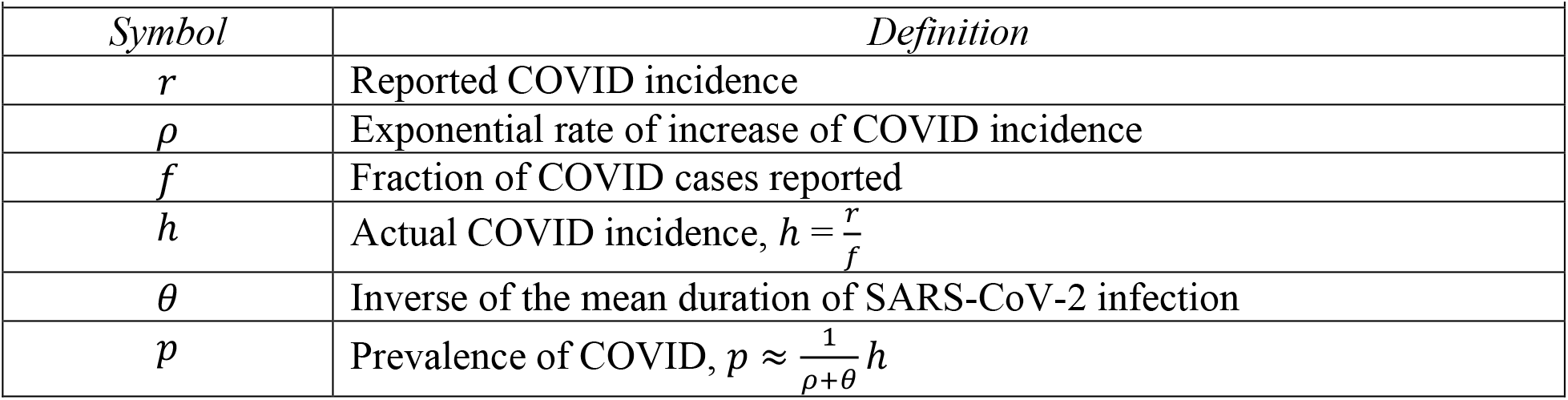
Component Parameters in the Derivation of COVID Prevalence, *p*.

### Data: Cohort of 250 High-COVID-Volume Hospitals

From a database of U.S. hospitals maintained by the Department of Health and Human services (HHS) [26], we identified the 250 hospitals with the highest cumulative volume of emergency department visits for COVID-19 from the week ending June 25, 2021 through the week ending December 10, 2021. These high-volume hospitals occupied 164 counties in 41 states and territories. Fig. 1 displays a map of hospital locations within the continental United States.

**Fig. 1.**
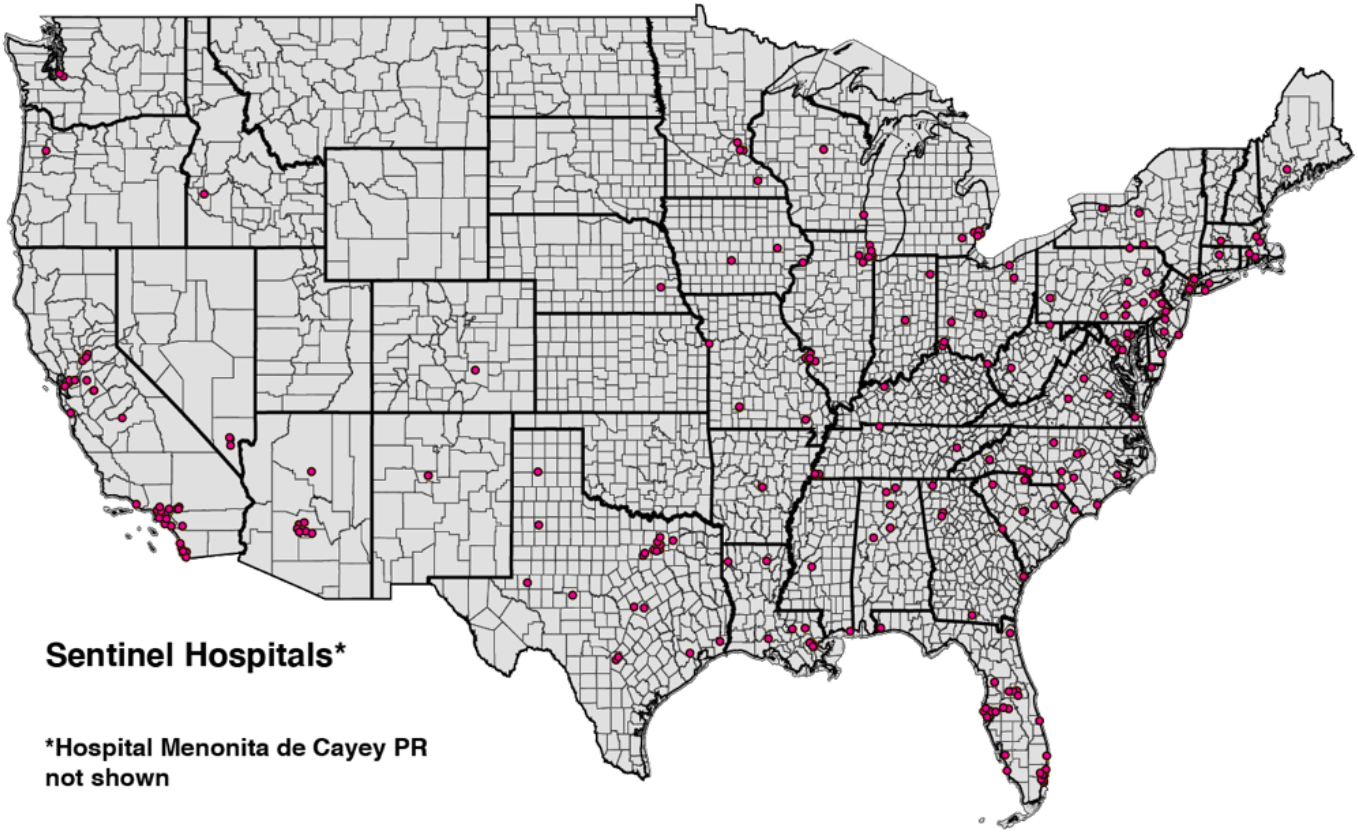
U.S. Continental Map Showing Locations of 249 of the 250 High-Volume Hospitals. Hospital Menonita de Cayey, Cayey, Puerto Rico, is not shown. State and county boundaries are indicated.

The hospital database maintained by HHS reported data on a weekly basis. For each of the 250 high-volume hospitals, we extracted the following three variables from the week ending June 25, 2021, through the week ending January 7, 2022.

1. *inpatient_beds_used_7_day_avg*, defined as “Average of total number of staffed inpatient beds that are occupied reported during the 7-day period.”
2. *total_adult_patients_hospitalized_confirmed_covid_7_day_avg*, defined as “Average number of patients currently hospitalized in an adult inpatient bed who have laboratory-confirmed COVID-19, including those in observation beds. This average includes patients who have both laboratory-confirmed COVID-19 and laboratory-confirmed influenza.”
3. *total_pediatric_patients_hospitalized_confirmed_and_suspected_covid_7_day_avg*, defined as “Average number of patients currently hospitalized in a pediatric inpatient bed, including NICU, PICU, newborn, and nursery, who are suspected or laboratory-confirmed-positive for COVID-19. This average includes those in observation beds reported in the 7-day period.”

### Estimating c, the Fraction of Hospitalized Patients Who Are COVID-Positive

For each hospital and each week, we calculated the number of COVID-19-positive patients as the sum of variables (ii) and (iii) above, while variable (i) gave the total number of hospital patients. For each week, summing over all 250 hospitals, we computed cohort-wide numbers of COVID-19-positive inpatients and total inpatients, from which we then computed *c*, the fraction of all inpatients who were COVID-19-positive. While we display the entire timeline of the fraction *c* in Fig. 2 below, we relied on the most recently available value of *c* for the week ending January 7, 2022, in our calculations of the fraction of incidental hospitalizations *π*.

**Fig. 2.**
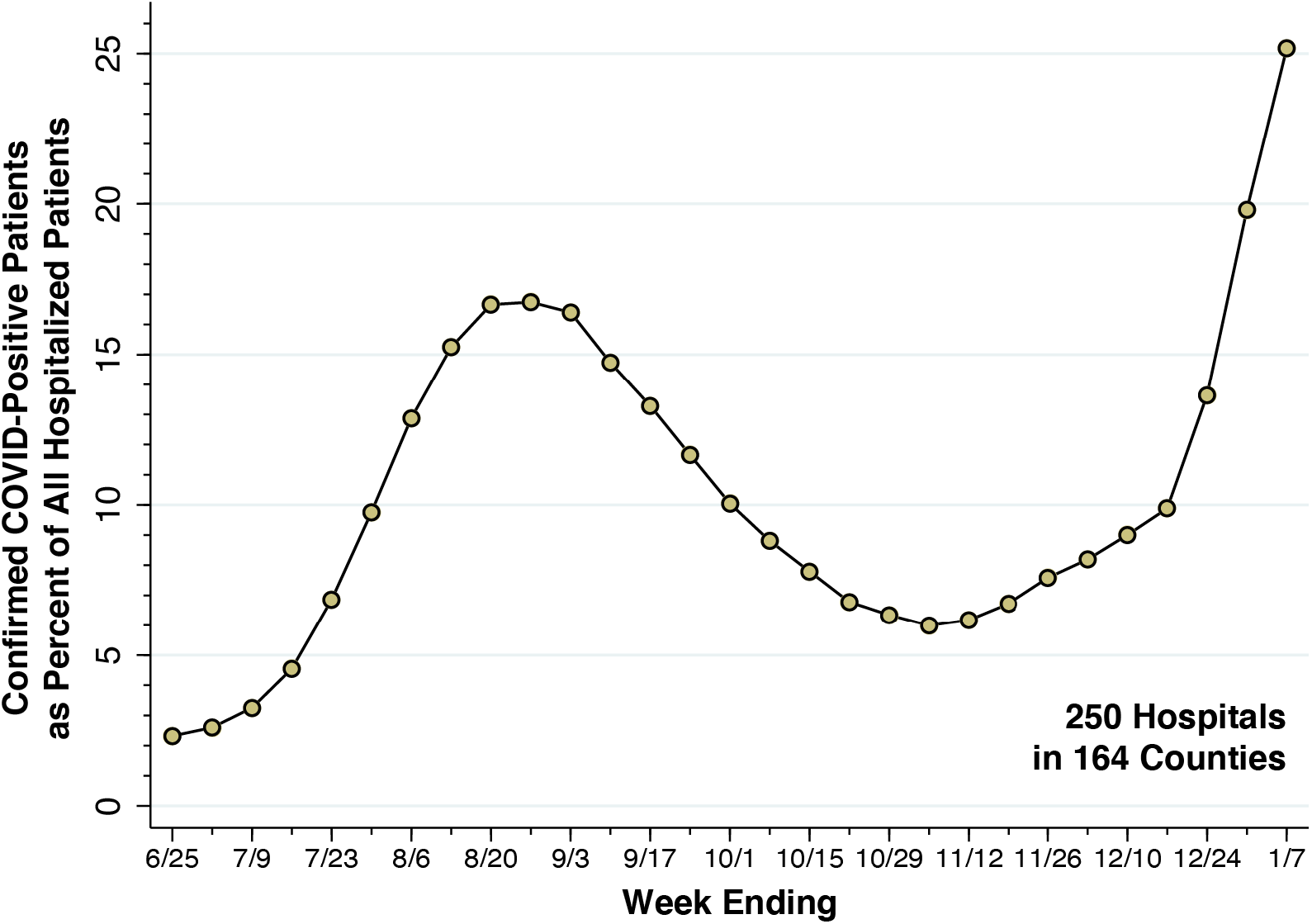
Fraction of All Inpatients Who Were COVID-Positive in a Cohort of 250 High-Volume Hospitals, Weeks Ending June 25, 2021, through January 7, 2022. Source: U.S. Department of Health and Human Services [26]. For the most recent week ending January 7, 2022, the fraction of inpatients who were COVID-positive was 0.2517, which we took as the value of the parameter *c*. The corresponding odds ratio was 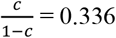. The underlying data are given in Supplement Table A1.

### Data: Confirmed COVID Incidence in 164 Counties

For each of the 164 counties covering our cohort of 250 high-volume hospitals, we downloaded weekly reported COVID-19 incidence from the *Counties* tab of Excel spreadsheets regularly issued by the White House COVID-19 Team as Community Profile Reports [27]. We extracted three variables from each report: *Population, Cases - last 7 days*, and *Cases per 100k - last 7 days*. These variables were extracted from the reports of December 20 (covering the week from December 13-19), December 27 (covering December 20-26), January 3 (covering December 27 – January 2), and January 10 (covering January 3-9). The resulting database contained a balanced panel of 4 serial observations on each of 164 counties. We show a whiskers-on-box plot of the variable *Cases per 100k - last 7 days* in Fig. 3 below.

**Fig. 3.**
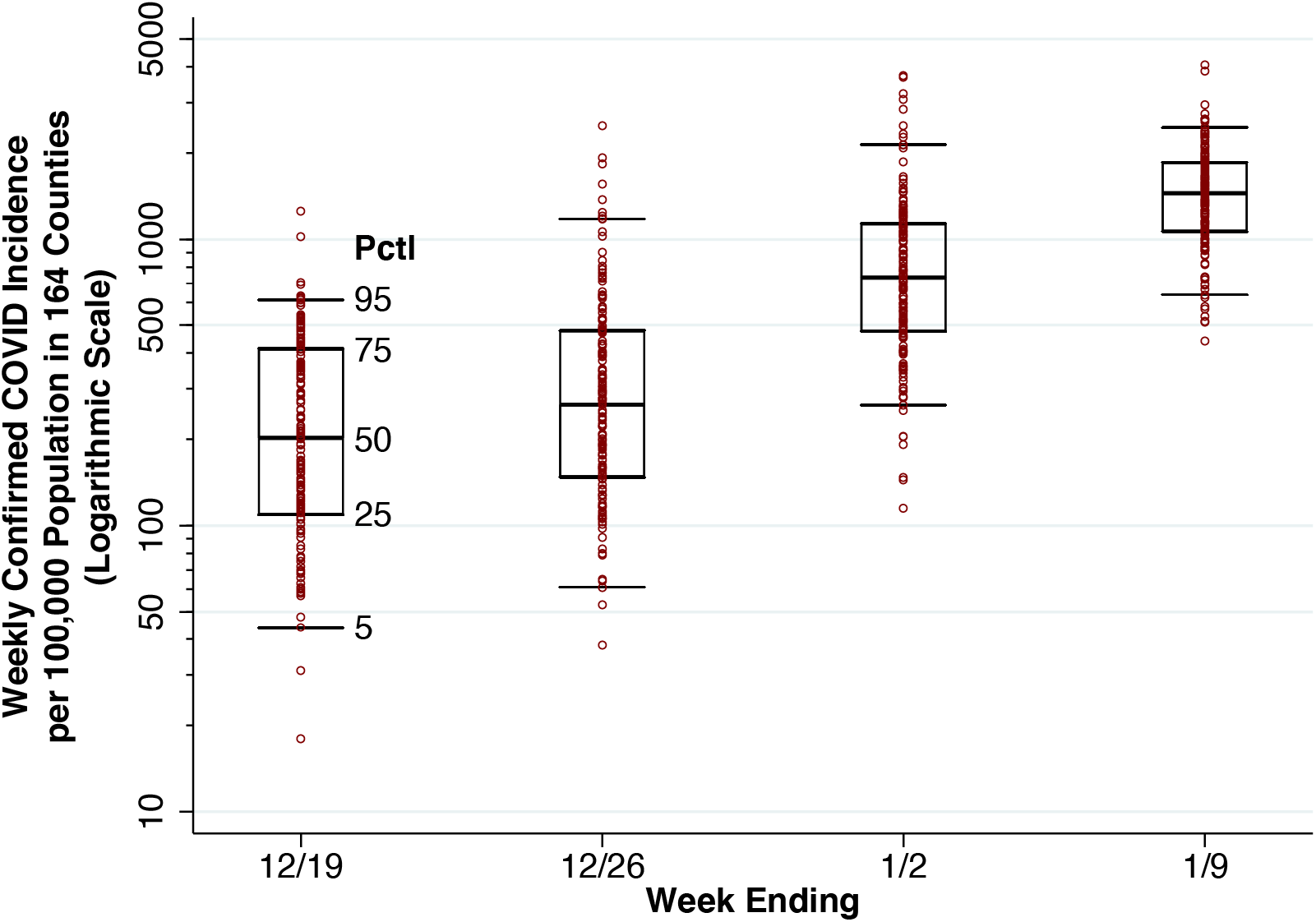
Whiskers-on-Box Plots of Weekly Confirmed COVID-19 Incidence per 100,000 Population in the 164 Counties Containing the 250 Study Hospitals, Weeks Ending December 19, 2021, Through January 9, 2022. For each week, the 5^th^, 25^th^, 50^th^, 75^th^, and 95^th^ percentiles are superimposed upon the individual county-specific datapoints. A total of 11 datapoints with zero incident cases are omitted from the first two weeks.

### Estimating the Exponential Rate of Increase of COVID Incidence, ρ, and Reported COVID Incidence, r_t_

We relied upon the county-specific data derived from the Community Profile Reports [27] to estimate the exponential rate of increase of COVID-19 incidence, *ρ*. To that end, we ran the following log-linear fixed-effects regression model on our panel of 4 serial weekly observations (indexed *t* = 1,…,4) on 164 counties (indexed *i* = 1,…,164):

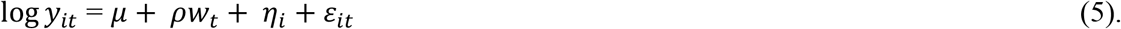

In equation (5), the observations *y*_*it*_ corresponded to the data variable *Cases - last 7 days*, while the observations *w*_*t*_ represented the ending date of each of the four weeks. The parameter *μ* was an overall constant term, the parameters *η*_*i*_ were county-specific fixed effects, and the terms *ε*_*it*_ were spherical errors. The parameter *ρ* was estimated by ordinary least squares.

To test whether the data adequately fit an exponential growth model, we plotted the reported incidence *r*_*t*_ for all 164 counties combined against *w*_*t*_ to check for serial correlation of residuals. The reported incidence was computed as *r*_*t*_ = ∑_*i*_ *n*_*i*_*m*_*it*_/∑_*i*_ *n*_*i*_, where *m*_*it*_ denotes *Cases per 100k - last 7 days* and *n*_*i*_ denotes *Population*. We show a plot of *r*_*t*_ versus *w*_*t*_ in Fig. 4 below. While we used a continuous-time model to develop our formula for prevalence in equation (3), here we use the discrete time notation to refer to computations made from the panel of 164 observations in each of 4 successive weeks.

**Fig. 4.**
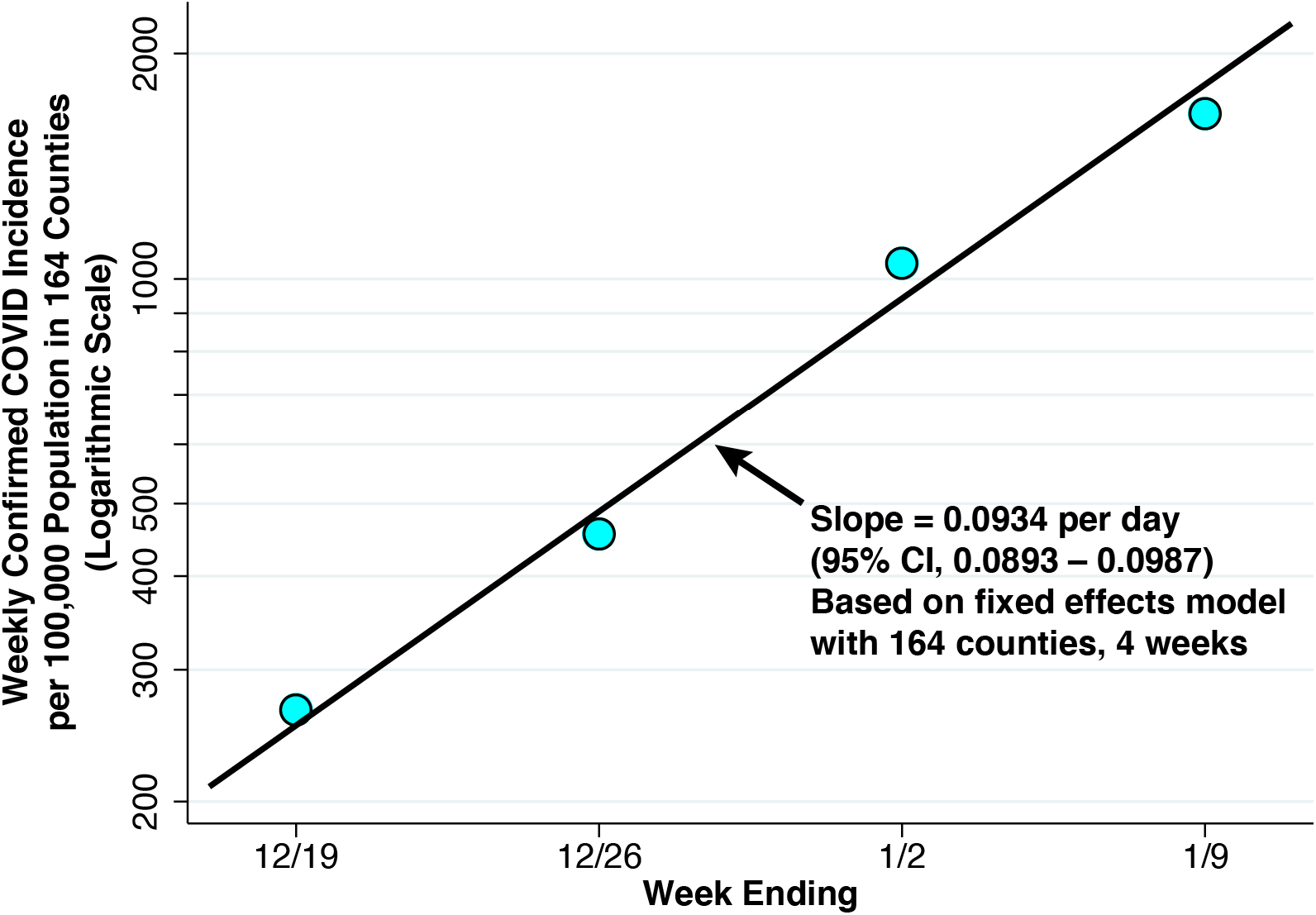
Weekly Confirmed COVID-19 Incidence per 100,000 Population in the 164 Counties Containing the 250 Study Hospitals, Weeks Ending December 19, 2021, Through January 9, 2022. The superimposed line has a slope of 0.0934 per day, derived from a linear fixed effects regression model of the logarithm of weekly COVID-19 cases versus the date on which each week ended (equation 5). The balanced panel had 4 observations for each of 164 counties. No serial correlation of residuals is evident.

### Estimating the Fraction of COVID-19 Cases Reported, f_t_, and Actual COVID-19 Incidence, h_t_

For our estimate of the fraction of COVID-19 cases reported, *f*_*t*_, we relied upon estimates issued by the Institute for Health Metrics and Evaluation (IHME) [28]. The authors estimated that by late December 2021 and early January 2022, *f*_*t*_, what they termed the “infection-detection rate,” had fallen to 0.25. (See Figure 8-1 in [28].) By relying on reported COVID-19 incidence *r*_*t*_ rather than actual COVID-19 incidence *h*_*t*_ to estimate *ρ* above, we effectively assumed *f*_*t*_ to be constant during the 4 weeks covered by our panel, and we do so here as well. Thus, for each week *t* = 1,…,4, we compute actual incidence as *h*_*t*_ = *r*_*t*_/0.25 = 4*r*_*t*_.

### Estimating q, the Proportion Hospitalized

A contemporaneous technical report from the UK Health Security Agency [29] noted that 3,019 Omicron cases were hospitalized among 528,176 Omicron cases total, which gives *q* ≈ 0.006. While hospitalization practices may differ in the UK’s National Health Service, this source has the advantage that the denominator closely approximated all Omicron infections, and not just symptomatic cases.

### Aggregate and County-Specific Estimates of the Fraction of Incidental Hospitalizations, π

We estimated the fraction of incidental hospitalizations *π* not only for all 164 counties combined, but also for each county individually. We focused on the first week in January, that is, for the final week *t* = 4 in our four-week panel. For the aggregate calculation, we estimated the reported incidence *r* and the proportion of hospitalized patients who are COVID-19-positive *c* for all counties combined. (Since we are focusing on a specific week, we have dropped the subscripts *t*.) Given the estimated parameters *ρ, f, θ*, and *q*, we then computed an aggregate value of *π*. For the county-specific calculations, we estimated the reported incidence *r*_*i*_ and the proportion of hospitalized patients who were COVID-positive *c*_*i*_ for each county *i* (again dropping the subscript *t*). Assuming the parameters *ρ, f, θ*, and *q* to be constant across counties, we then computed county-specific values for each *π*_*i*_.

## Results

### Aggregate Fraction of Hospitalized Patients Who Are COVID-19-Positive, c

From the week ending June 25, 2021, through the week ending January 7, 2022, Fig. 2 plots the fraction of all hospital inpatients who were COVID-19-positive in our cohort of 250 high-volume hospitals. During the summer’s Delta wave, this fraction rose to 16.74 percent for the week ending August 27, 2021. During the Omicron wave, the fraction rose rapidly rising to 25.17 percent for the week ending January 7, 2022. We took the latter datum as our estimate of the parameter *c*. Accordingly, about one in four inpatients of our cohort of 250 high-COVID-volume hospitals was hospitalized.

### Growth Rate of COVID-19 Incidence, 164 Counties

Fig. 3 displays a whiskers-on-box plot of reported weekly COVID-19 incidence *r*_*it*_ in the 164 counties containing our 250-hospital cohort. The plot covers the four weeks of data through the week ending January 9, 2022. During the latter week, Potter County TX, which includes Amarillo, was at the 25^th^ percentile of reported incidence (1,069 cases per 100,000 population). Baltimore County MD was at the median (1,466 cases per 100,000 population), and Hartford County CT was at the 75^th^ percentile (1,858 cases per 100,000 population). The two counties with the highest reported incidence were Miami-Dade County FL (4,065 per 100,000 population) and Bronx County NY (3,865 per 100,000 population).

For the same four weeks, Fig. 4 plots the population-weighted mean values of confirmed COVID-19 incidence among all 164 counties combined. These values represent our estimates of reported incidence *r*_*t*_. The fixed-effect log linear regression described on our panel of 164 counties over 4 weeks, described in equation (5), gave an estimate of *ρ* = 0.0934 per day (95% confidence interval: 0.0893 – 0.0987). The fitted line in the figure with the same slope shows no evidence of serial correlation of residuals, thus supporting a model of recent exponential growth.

### Actual COVID-19 Incidence and Prevalence, 164 Counties

For the most recent week ending January 9, 2022, the mean weekly confirmed COVID-19 incidence for the 164 counties in our panel was 1,663 per 100,000 population (Fig. 3), which comes to a reported incidence of *r* = 237.6 per 100,000 per day. Given our estimate of the fraction of cases reported at *f* = 0.25, we obtain an estimate of actual incidence of *h* = *r*/*f* = 950.4 per 100,000 per day.

The U.S. Centers for Disease Control and Prevention (CDC) has estimated that the mean duration of infectiousness for Omicron is 5-6 days [30]. That gives us *θ* = 1/5.5 = 0.182. From equation (3), we thus estimate actual COVID prevalence in our panel of 164 counties during the most recent week ending January 9 to be 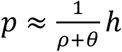, which comes to 3.45 percent of the population.

### Estimated Fraction of Incidental Hospitalizations

Given estimates of the parameters *q* = 0.006, *c* = 0.2517, and *p* = 0.0345, we relied upon equation (2) to compute 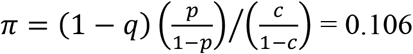. That is, we estimate 10.6 percent incidental COVID hospitalizations for the 164 counties containing our high-volume hospital cohort during the first week of January 2022. Our calculations are summarized in Table 3.

**Table 3.**
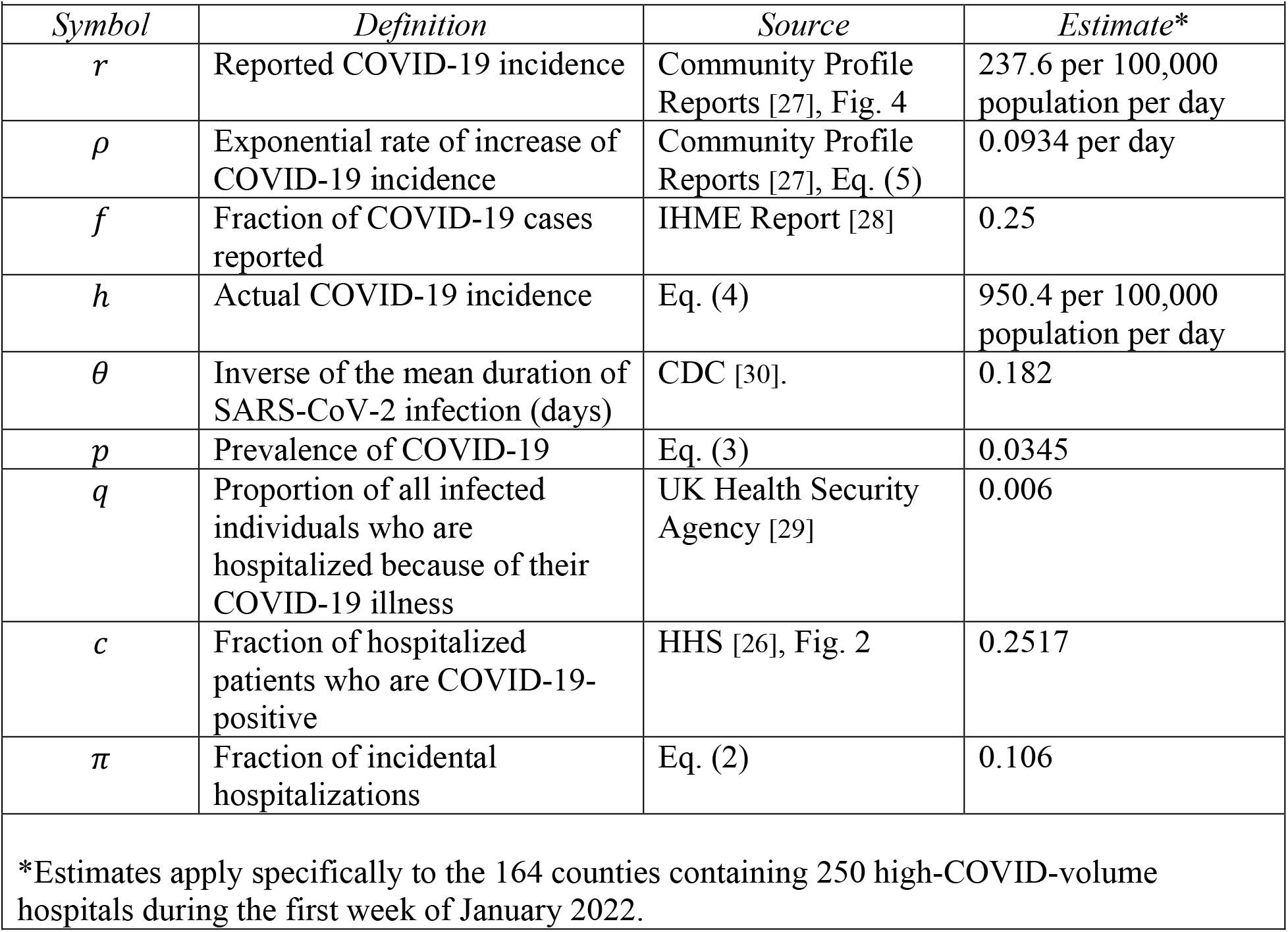
Estimates of Component Parameters in the Derivation of the Fraction of Incidental Hospitalizations, *π*.

### Variability of the Fraction of Incidental Hospitalizations, π, Across Counties

Our overall estimate of *π* = 10.6 percent is an average that does not capture its variability across the 164 counties under study. When we applied the formula of equation (2) individually to each county, once again restricting the computations to the first week of January 2022, the median value of *π* was 9.5 percent, with an interquartile range of 6.7 to 12.7 percent.

Fig. 5 displays this variability in the form of a scatterplot of 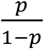 versus 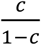 for 162 of 164 counties. If all counties had the same value of *π*, they would line up along the superimposed ray from the origin. Those datapoints above the ray have higher values of *π*, while those below the ray have lower values. Geometrically speaking, the value of *π* at each datapoint depends on the slope of the ray drawn from that point back to the origin. Thus, Allegheny County PA, which houses Pittsburgh, has the highest estimated value of 40.4 percent. By contrast, while Miami-Dade has the highest estimated prevalence of *p* = 8.4 percent, the estimated fraction of incidental COVID hospitalizations is 14.1 percent. As equation (2) shows, when the overall prevalence *p* of COVID-19 in the county is higher, the fraction of incidental COVID-19 hospitalizations *π* tends to be higher. But as hospitals in the county fill up with COVID-19 patients, thus pushing *c* higher, the fraction of incidental COVID hospitalizations *π* tends to go lower.

**Fig. 5.**
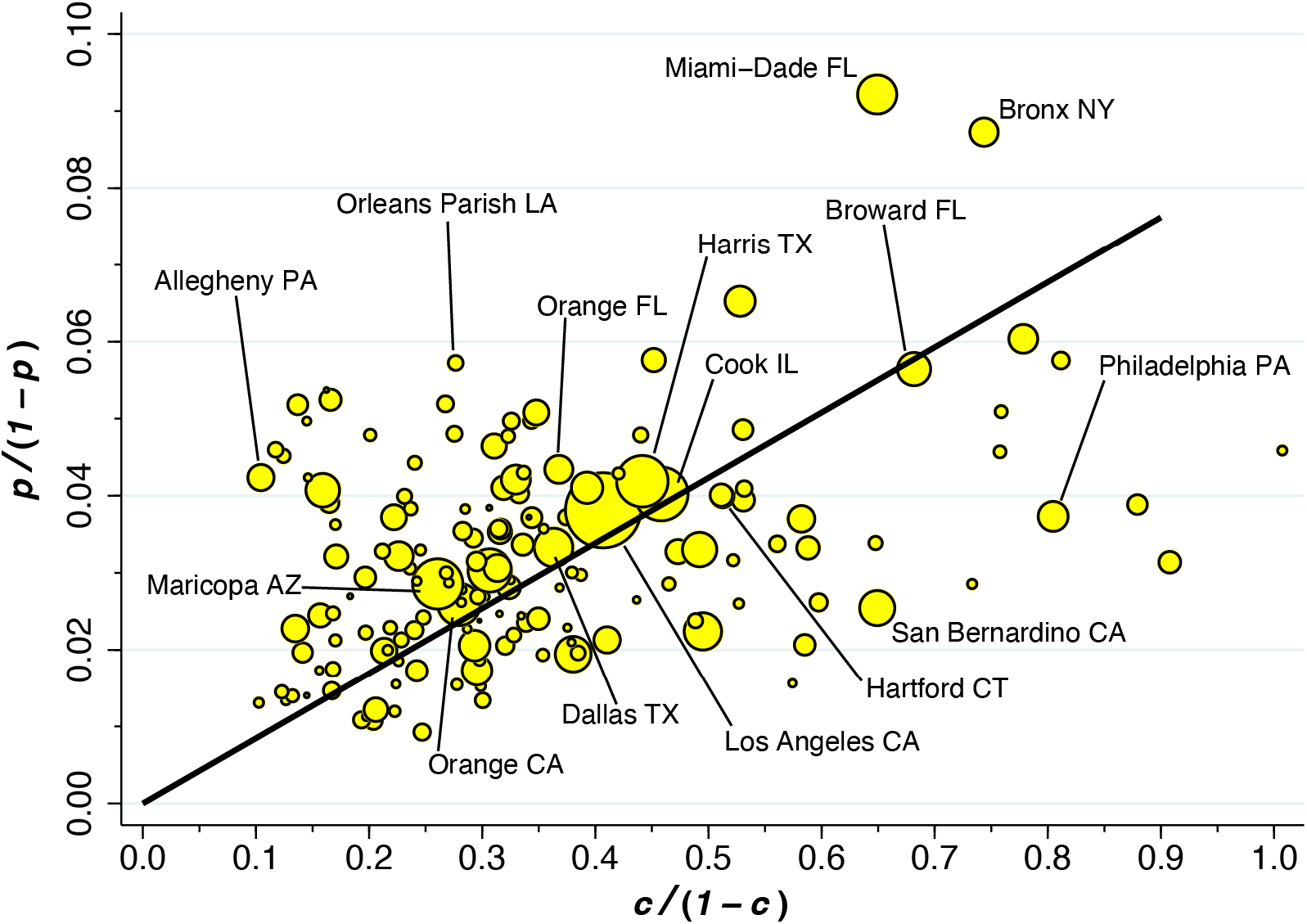
Plot of *p*/(1 − *p*) Versus *c*/(1 − *c*) Among 162 or 164 Counties During the First Week of January 2022. The ray from the origin has slope 0.085 (95% CI: 0.079 – 0.090) based on a population-weighted regression. *p* = prevalence of COVID; *c* = fraction of hospitalized patients who are COVID-positive.

## Discussion

Assuming only one-quarter (that is, *f* = 0.25) of all Omicron infections were reported by public authorities, we estimated the aggregate prevalence of active SARS-CoV-2 infection during the first week of January to be 3.45% (that is, *p* = 0.0345). During the same week, among 250 high-COVID-volume hospitals within our 164-county panel, an estimated 1 in 4 inpatients were found to be COVID-19-positive (more precisely, *c* = 0.2517). Applying these estimates to our population-based formula, we found that among all COVID-19-positive hospitalized patients in all 164 counties combined, an estimated 10.6% were incidental infections (that is, *π* = 0.106). Across individual counties, the median fraction of incidental COVID-19 hospitalizations was 9.5%, with an interquartile range of 6.7 to 12.7%.

### Limitations

The fundamental drawback of the current study is that it is a modeling exercise. It does not adhere to the standard of a prospective study of hospitalized COVID-19 patients set forth in the Introduction. As a modeling exercise, its principal limitations include uncertainty in the fraction *f* of reported cases; its critical but non-testable assumption about equality of hospitalization rates; its reliance on non-U.S. data sources for assessing the probability *q* that a SARS-CoV-2 infected individual would have a severe case requiring hospitalization; and the significant variability in the estimated fraction *π* of incidental cases across counties. Each of these limitations is addressed in turn.

### Fraction of Reported COVID-19 Cases, f, as a Principal Source of Uncertainty

The principal source of uncertainty in our estimate of incidental COVID-19 hospitalization is the fraction *f* of reported COVID cases. During the Omicron wave, as noted above, this fraction declined to 25 percent of actual cases [28], in great part as a result of a surge in asymptomatic and mildly symptomatic infections, as well as an increasing proportion of rapid test results not tabulated by public health authorities. Our formula for the prevalence *p* of infection (equation 3) is a linear function of the actual incidence *h*, which is turn directly proportional to the inverse of *f* (equation 4). Thus, if only 20 percent of cases were reported (that is, *f* = 0.20), our estimate of COVID-19 prevalence *p* would increase to 4.3 percent and the resulting estimate of the fraction of incidental hospitalizations *π* would rise to 13.3 percent. At the other extreme, if as many as 50 percent of cases were reported (that is, *f* = 0.50), our estimate of COVID-19 prevalence *p* would drop to 1.39 percent and the resulting estimate of *π* would fall to 4.1 percent.

### Equality of Hospitalization Rates as a Critical Assumption

In the derivation of our population-based formula, we made the key assumption that those individuals without COVID-19 have the same probability *g* of hospitalization as those with non-severe COVID-19. Strict adherence to the notion of causality would seem to require this assumption. If a concurrent COVID-19 infection increases the probability of hospitalization, then the infection has a causal role in the hospitalization and is thus not incidental. If a hospitalization is purely incidental, then the probability of hospitalization would be the same with or without COVID-19.

Consider again our patient the hypertensive heart disease who, as a result of high fevers and dehydration from an Omicron infection, develops atrial fibrillation, a cardiac arrhythmia. He needs to be hospitalized to get his heart rate down and convert his heart rhythm back to normal. His would not be an incidental COVID-19 hospitalization. Consider instead a patient with a history of extreme myopia since childhood who suffers a spontaneous retinal detachment and is hospitalized for eye surgery. During her admission workup, she is found to be COVID-19-positive. Since the infection did not apparently affect her probability of hospitalization, hers would be an incidental COVID-19 hospitalization. (This is not to deny that the COVID-19 epidemic has delayed the time when patients have sought treatment and thus affected the severity of vision loss upon initial presentation [31].)

The problem with the foregoing logic is that individuals in the two groups – those without COVID-19 and those with non-severe COVID-19 – are different people. Thus, one individual with a white-collar occupation may be able to work remotely and does not contract COVID-19. Another individual with a blue-collar occupation cannot work remotely and comes down with a non-severe infection at work. The latter individual may also have a higher risk of hospitalization from an onsite workplace accident.

Generalizing our formula for *π*, we suppose that those without a COVID-19 infection have a hospitalization probability *g*′ that is not necessarily equal to the hospitalization probability *g* of those with non-severe COVID-19. Then the fraction of incidental COVID-19 hospitalizations becomes *Rπ*, where 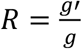. is the relative risk of hospitalization between the two groups and *π* is as already defined in equation (2). This does not mean that our strong assumption that *R* = 1 necessarily understates *π*. In the foregoing example comparing white-collar and blue-collar workers, *R* < 1 and thus *π* would be overstated. More generally, when those individuals who take precautions to reduce the risk of infection, such as getting vaccinated [32], also tend to adopt other preventive measures to reduce the risk of hospitalization generally, such as not smoking, our strong assumption that *R* = 1 will tend to overstate *π*.

### Reliability of the Estimate of q from UK Data

Relying on data reported by the UK Health Security Agency [29], we took *q*, the probability that an infected individual would have a severe case requiring hospitalization, as 0.006. It is worth inquiring whether there are any U.S.-based sources that might provide a more reliable estimate. The difficulty is that computation of *q* needs to be based upon a population denominator that includes all cases of COVID-19, even asymptomatic and unreported cases.

A study of symptomatic patients infected with the Omicron variant in the Houston Methodist hospital system [33] revealed that out of 2,232 symptomatic patients, 313 were admitted to the hospital. This source, it would seem, yields an estimate of *q* = 0.14, which is an order of magnitude greater than the estimate derived from the UK data. The problem with relying upon this alternative data source is that the denominator reflects symptomatic patients primarily presenting to the emergency department, rather than all COVID-19 cases. In fact, the percentage reported by Houston Methodist is quite close to the rate of 15.1 hospital admissions per 100 emergency department visits for COVID that we computed from our 250-hospital cohort, as shown in Supplement Fig. A1.

### Variability of π Across Counties

While our aggregate measure of the incidental COVID-19 fraction for all 164 counties is on the order of 11 percent, Fig. 5 shows significant variability across individual counties. Some of this variability is no doubt due to sampling errors in the proportions *c*_*i*_ of inpatients who were COVID-19-positive, especially in counties with only one high-volume hospital. Our assumption that the fraction of COVID-19 cases reported *f* was uniform across counties may have introduced errors in the county-specific incidence estimates *h*_*i*_ = *r*_*i*_/*f*. Likewise, our assumption that the exponential rate of increase of COVID-19 incidence *ρ* was uniform across counties may have introduced errors in the county-specific prevalence estimates 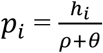.

Still, some of the variability of the variability may be due to systemic differences between hospitals in their range of services and patient populations. For example, a hospital may have a large, specialized transplant service with many immunosuppressed patients who are persistently COVID-19-positive.

### Extensions

We chose to analyze the time period from December 2021 to January 2022 when COVID-19 incidence was exponential rising (Figs. 3 and 4). This choice allowed us to apply a simplified formula (equation (3)) to estimate the prevalence *p* of the disease. We focused on the Omicron BA.1 wave, when the reported incidence of infection was markedly higher than in other waves thus far observed in the United States. During such a period of maximum disease prevalence, we would expect the fraction *π* of incidental COVID-19 hospitalizations to likewise have approached its peak (equation (2)). Still, our methodology can be extended to other time periods during the SARS-CoV-2 epidemic. Application to extended intervals when incidence is neither uniformly increasing nor decreasing would require a more complex model of disease prevalence, as derived in the Supplement.

We chose to analyze a longitudinal panel of 164 counties throughout the United States. This choice was intended to cover a representative, high-density, mostly urban population. Still, our methodology can be extended to other countries and to population subgroups based on demographic characteristics, occupation, and vaccination status. Such extensions would require group-specific data on the prevalence *p* of infection, which would in turn require group-specific estimates of the fraction *f* underreported. We would also need group-specific data on the fraction *c* of all hospitalized patients who are COVID-19 positive as well as the proportion *q* of infected individuals who are hospitalized because of their COVID-19 illness.

### Policy Implications

Our findings imply that, in the aggregate, the burden of patients admitted for complications of their COVID-19 infections is far greater than the number of incidental cases. Consequently, real-time surveillance data on COVID-19 hospitalization rates can still be relied upon to gauge the clinical severity of the disease as well as its impact on limited healthcare manpower [34,35], other limited healthcare resources [36–38], healthcare costs [39], and excess mortality [40].

Moreover, COVID-19 hospitalization rates can still be used as a reliable endpoint to evaluate public policies to reduce morbidity and mortality from the disease. In particular, the repeated observation [32,41] that populations with higher rates of vaccination against SARS-CoV-2 have lower risks of hospitalization similarly retains its public policy significance.

Hospitalization rates can likewise continue to be used as an endpoint to assess the impact on non-pharmacological public policies [42].

## Conclusion

Our population-based estimates suggest that incidental COVID-19 infections have indeed been a nontrivial fraction of all COVID-positive hospitalized patients, but they fall far short of the proportions suggested by some sources [2-4,7,10,14,15]. Even under our conservative assumption that there were four times as many COVID-19 infections as cases reported by public authorities, there were not nearly enough prevalent COVID-19 cases to push the fraction of incidental hospitalizations even close to the one-half mark suggested by those sources. Our examples of the patient with hypertensive heart disease who developed a cardiac arrythmia and the young man who got into a car crash as a result of their viral infections suggest that the indirect methodologies employed in prior studies have been overly restrictive in their assessment of COVID-19-caused admissions.

## Data Availability

Data and analyses are available via the Open Science Framework (OSF) at https://osf.io/7umje/

