## Supplement for "Population-Based Model of the Fraction of Incidental COVID-19 Hospitalizations During the Omicron BA.1 Wave in the United States"

*Derivation of Equation (2) in the Main Text*

Let $p$ (where 1 > $p$ > 0) denote the proportion of individuals in the entire population who are COVID-19 cases, that is, the *prevalence* of COVID-19. Let $q$ (where 1 > $q$ > 0) denote the proportion of all COVID-19 cases that are *severe*, that is, hospitalized *because* of their illness. We make the key assumption that those individuals without COVID-19 have the same probability $g$ of hospitalization (where 1 > $g$ > 0) as those with non-severe COVID-19. The proportion of individuals in the population who do not have COVID-19 *and* are hospitalized is thus $g(1-p)$. The proportion who have non-severe COVID-19 *and* are incidentally hospitalized as $g(1-q)p$, while the proportion who have severe COVID-19 *and* are hospitalized as $qp$.

Let $c$ (where 1 > $c$ > 0) denote the fraction of all hospitalized individuals who are COVID-19-positive, whether those individuals are severe or non-severe cases. Utilizing the foregoing expressions, we have $c = \frac{g(1-q)p+qp}{g(1-p)+(1-q)p+qp}$. We can write the odds that a hospitalized patient is COVID-positive as $\frac{c}{1-c} = \frac{g(1-q)p+qp}{g(1-p)}$, which can be rewritten as $\left(\frac{p(1-q)}{1-p}\right)\left(\frac{g(1-q)+q}{g\,(1-q)}\right)$. Utilizing the definition of $\pi$ in equation (1) in the main text and rearranging terms gives:

$$\pi = (1-q)\left(\frac{p}{1-p}\right)\Big/\left(\frac{c}{1-c}\right) \qquad (2).$$

*Derivation of Equation (3) in the Main Text*

Let $h(t)$ denote the incidence rate of SARS-CoV-2 infection at time $t \geq 0$. We assume that incidence is growing exponentially, that is, $h(t) = h_0 e^{\rho t}$, where $h_0, \rho > 0$. Let the duration $s > 0$ of infection have an exponential distribution with mean $1/\theta$, where $\theta$ > 0, so that the cumulative distribution function of $s$ is $F(s) = 1 - e^{-\theta s}$ and the corresponding survival function is $G(s) = 1 - F(s) = e^{-\theta s}$. Given these functional forms for $h$ and $G$, the prevalence of COVID-19 at time $t$ is $p(t) = \int_0^t h(u)G(t-u)\,du = \frac{h_0}{\rho+\theta}\left(e^{\rho t} - e^{-\theta t}\right)$. For sufficiently large $t$, the second term inside the parentheses gets small, so that we have:

$$p(t) \approx \frac{1}{\rho+\theta} h(t) \qquad (3).$$

*Tables*

**Table A1. Data Underlying Figure 2.** [a]

| *Week Ending* | *Total Hospital Inpatients* | *COVID-19 Positive Inpatients* | *Percent COVID-19 Positive (c)* |
|---|---|---|---|
| 6/25/21 | 103,859 | 2,409 | 2.32 |
| 7/2/21 | 102,030 | 2,652 | 2.60 |
| 7/9/21 | 105,387 | 3,426 | 3.25 |
| 7/16/21 | 106,785 | 4,857 | 4.55 |
| 7/23/21 | 107,265 | 7,354 | 6.86 |
| 7/30/21 | 107,556 | 10,500 | 9.76 |
| 8/6/21 | 107,758 | 13,875 | 12.88 |
| 8/13/21 | 108,483 | 16,537 | 15.24 |
| 8/20/21 | 108,509 | 18,079 | 16.66 |
| 8/27/21 | 108,193 | 18,115 | 16.74 |
| 9/3/21 | 107,151 | 17,575 | 16.40 |
| 9/10/21 | 109,515 | 16,130 | 14.73 |
| 9/17/21 | 108,878 | 14,470 | 13.29 |
| 9/24/21 | 108,083 | 12,598 | 11.66 |
| 10/1/21 | 107,455 | 10,793 | 10.04 |
| 10/8/21 | 107,605 | 9,480 | 8.81 |
| 10/15/21 | 107,123 | 8,348 | 7.79 |
| 10/22/21 | 107,114 | 7,261 | 6.78 |
| 10/29/21 | 106,776 | 6,774 | 6.34 |
| 11/5/21 | 107,227 | 6,426 | 5.99 |
| 11/12/21 | 107,658 | 6,646 | 6.17 |
| 11/19/21 | 105,426 | 7,083 | 6.72 |
| 11/26/21 | 104,164 | 7,897 | 7.58 |
| 12/3/21 | 109,395 | 8,962 | 8.19 |
| 12/10/21 | 109,638 | 9,873 | 9.01 |
| 12/17/21 | 107,399 | 10,616 | 9.88 |
| 12/24/21 | 101,122 | 13,797 | 13.64 |
| 12/31/21 | 106,676 | 21,126 | 19.80 |
| 1/7/22 | 109,725 | 27,621 | 25.17 |

a. Total Hospital Inpatients and COVID-19 Positive Inpatients are 7-day averages. $c = 100 \times$ (COVID-19 Positive) ÷ (Total Inpatients). Source: U.S. Department of Health and Human Services [1].

*Figures*

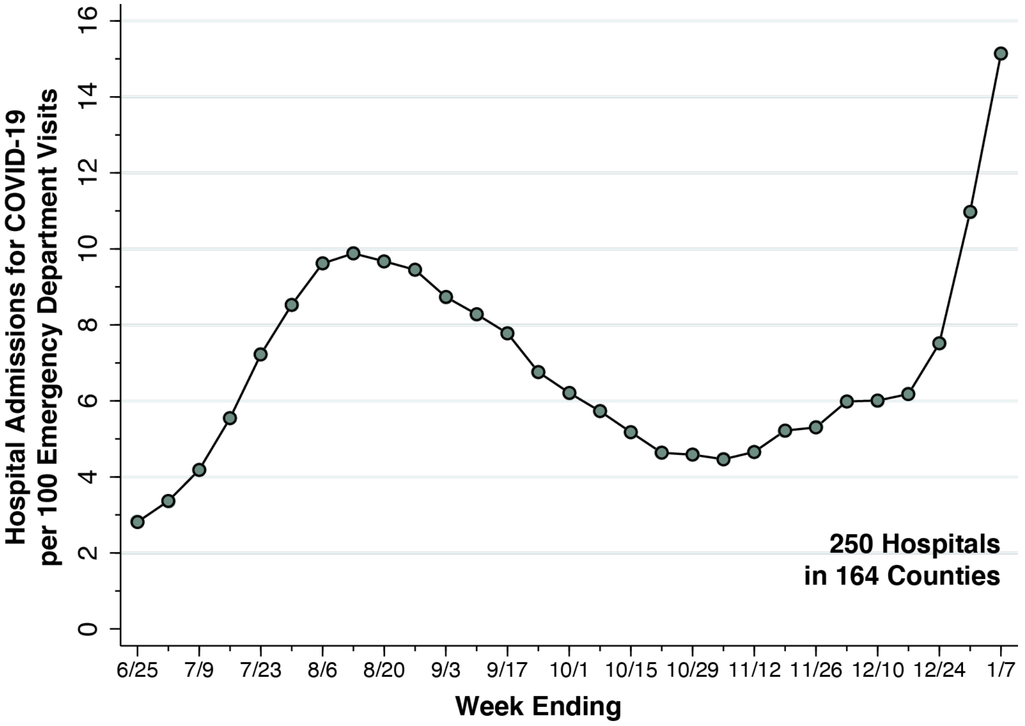


**Fig. A1. Hospital Admissions for COVID-19 per 100 Emergency Department Visits in a Cohort of 250 High-Volume Hospitals, Weeks Ending June 25, 2021, through January 7, 2022.** See Technical Notes to Fig. A1 below.

*Technical Notes to Fig. A1.*

The data source was U.S. Department of Health and Human Services [1], as described under *Data: Cohort of 250 High-Volume Hospitals* in the section on Methods and Data in the main text.

Emergency department visits to each of the cohort hospitals was derived from the variable *previous_day_covid_ED_visits_7_day_sum*, defined as "Sum of total number of ED visits who were seen on the previous calendar day who had a visit related to COVID-19 (meets suspected or confirmed definition or presents for COVID diagnostic testing – do not count patients who present for pre-procedure screening) reported in 7-day period."

Hospital admissions were determined as the sum of two variables:

a) *previous_day_admission_adult_covid_confirmed_7_day_sum*, defined as "Sum of number of patients who were admitted to an adult inpatient bed on the previous calendar day who had confirmed COVID-19 at the time of admission reported in the 7-day period."

b) *previous_day_admission_pediatric_covid_confirmed_7_day_sum*, defined as "Sum of number of pediatric patients who were admitted to an inpatient bed, including NICU, PICU, newborn, and nursery, on the previous calendar day who had confirmed COVID-19 at the time of admission."
